# The neurodevelopment of anomalous perception: Evidence in cortical folding patterns for prenatal predispositions to hallucinations in schizophrenia

**DOI:** 10.1101/2020.06.04.20122424

**Authors:** Colleen P.E. Rollins, Jane R. Garrison, Maite Arribas, Aida Seyedsalehi, Zhi Li, Raymond C.K. Chan, Junwei Yang, Duo Wang, Pietro Lio, Chao Yan, Zheng-hui Yi, Arnaud Cachia, Rachel Upthegrove, Bill Deakin, Jon S. Simons, Graham K. Murray, John Suckling

## Abstract

**Background:** All perception is a construction of the brain from sensory input. Our first perceptions begin during gestation, making fetal brain development fundamental to how we experience a diverse world. Hallucinations are percepts without origin in physical reality that occur in health and disease. Despite longstanding research on the brain structures supporting hallucinations and on perinatal contributions to the pathophysiology of schizophrenia, what links these two distinct lines of research remains unclear.

**Methods:** We studied two independent datasets of patients with schizophrenia who underwent clinical assessment and 3T structural magnetic resonance (MR) imaging from the United Kingdom and Shanghai, China (n = 181 combined) and 63 healthy controls from Shanghai. Participants were stratified into those with (n = 79 UK; n = 22 Shanghai) and without (n = 43 UK; n = 37 Shanghai) hallucinations from the PANSS P3 scores for hallucinatory behaviour. We quantified the length, depth, and asymmetry indices of the paracingulate and superior temporal sulci (PCS, STS) from MR images and constructed cortical folding covariance matrices organized by large-scale networks.

**Results:** In both ethnic groups, we replicated a significantly shorter left PCS in patients with hallucinations compared to those without, and healthy controls. Reduced PCS length and STS depth corresponded to focal deviations in their geometry and to significantly increased covariance within and between areas of the salience and auditory networks.

**Conclusion:** The discovery of neurodevelopmental alterations contributing to hallucinations establishes testable models for these enigmatic, sometimes highly distressing, perceptions and provides mechanistic insight into the pathological consequences of prenatal origins.

## Introduction

All perception is a construct of the brain. Yet occasionally, sensory constructions emerge without origin in the physical world and are experienced as hallucinations. Despite over 20 years of active neuroimaging research on hallucinations (1–3), the neural systems supporting anomalous perceptual experiences remain disputed. Hallucinations occur transdiagnosticallly, cross-culturally, and in all sensory modalities (4). Although multiple brain mechanisms likely exist to support the diversity of hallucinations (5), our prior work established the role of sulcal topology, a product of early neurodevelopment (6), in reality monitoring and the experience of hallucinations associated with schizophrenia (7, 8).

The characteristic morphological features of the brain’s surface emerge in a specific order during the perinatal period, starting with the primary involutions occurring in the third trimester (9, 10). Gyral and sulcal architecture is intrinsically related to the brain’s functional organization (11). Functional neuroimaging studies have consistently reported alterations in the brain’s resting state networks (RSN) in those who experience hallucinations, particularly in the salience network, which engages the anterior cingulate and anterior insula to determine the origin and salience of internal and external stimuli (12, 13). The salience network equally coordinates the transition between functional networks related to self- and task-processing, suggesting cross-network abnormalities in the manifestation of hallucinations (14). Covariance patterns of cortical structure recapitulate RSN topography and reflect developmental coordination between brain regions, giving insights into the emergence of functional connectivity and indexing network integrity (15).

The paracingulate sulcus (PCS) is a complex structure on the medial prefrontal cortical surface, found only in humans and chimpanzees (16), that lies dorsal to the cingulate sulcus. It is characterized by high inter-individual and inter-hemispheric variability, including fragmentation, intersection by other sulci, and even absence (Figure 1). The PCS shows a notable leftward asymmetry among the general population that is reduced in individuals with a diagnosis of schizophrenia (17–19). We have previously shown that the bilateral absence of the PCS is associated with impairments in reality monitoring in healthy individuals (7) and a shorter PCS is associated with a predisposition to hallucinations in a study of 113 patients with schizophrenia (8).

**Figure 1.**
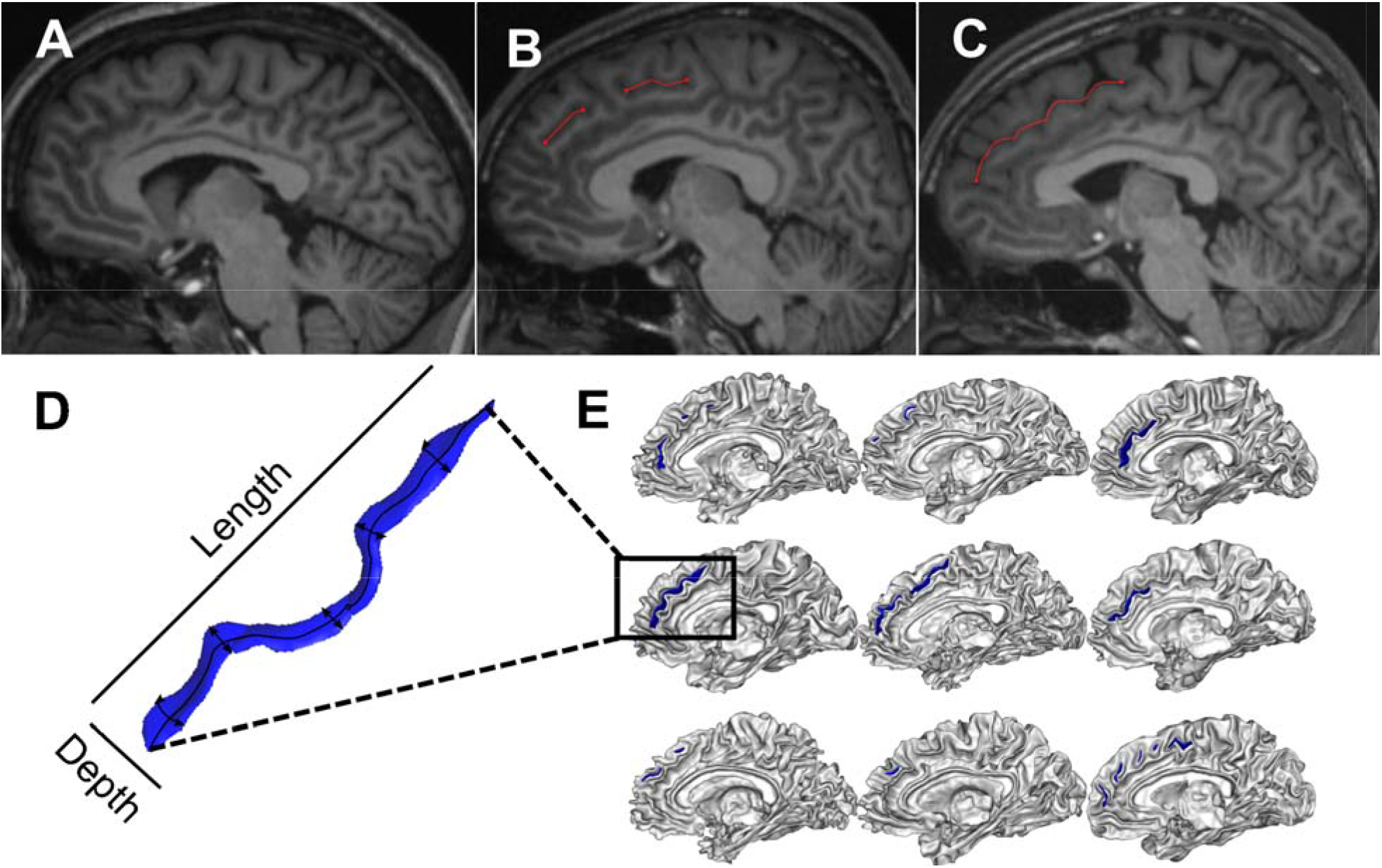
Variability in paracingulate sulcus (PCS) morphology. The PCS may be absent (A), fragmented by other sulci (B), or continuous (C). Sulci can be quantified through different shape measurements such as depth and length (D). Illustrative examples of PCS organization are shown), e n in (E) for n = 9 participants.

Sulcal contributions to hallucinations are not limited to the PCS. Localized deviations in the superior temporal sulcus (STS) in schizophrenia patients with auditory hallucinations have been reported (20, 21), as well as global decreases in cortical sulcation in patients with auditory and visual hallucinations (22). The PCS and STS overlap in their temporal emergence during sulcal ontogenesis at the fetal stage: the central sulcus forms around 19–25 weeks of gestation, with its secondary branches and the PCS appearing around 30 weeks (10) and the STS forms near 26 weeks, with the right STS emerging one week prior to that on the left (23). The STS displays a robust rightward depth asymmetry at the base of Heschl’s gyrus, regardless of language dominance and certain atypical conditions (23), though this asymmetry has not been investigated in schizophrenia. Due to the stability of sulci patterns during brain maturation (6), their topological variants in adulthood imply deviations in early brain development resulting in an intransient structural marker for the reality monitoring network, and a risk factor for experiencing hallucinations. This aligns with the developmental risk model of schizophrenia, which integrates perinatal hazards and neurodevelopmental processes along with postnatal experiences like urban upbringing or childhood trauma into the disorder’s pathogenesis (24).

Here we studied two ethnically independent structural MRI datasets of patients with schizophrenia (n = 181) and healthy controls (n = 63) to empirically test theoretical predictions linking hallucinations, cortical folding patterns, and salience and auditory brain network congruity. We first directly replicated, in this larger sample, the reduced left PCS length in schizophrenia patients with hallucinations compared to those without (8, 25), showing ethnic invariance (26) of a prenatally-determined structural marker for hallucinations. To render this tractable for large datasets, a semi-automated method was used to detect and characterize the PCS and STS from T1-weighted MRI, which we validated against the prior, gold standard manual approach. Sulcal 3D-segmentations allowed visualization of their geometry, illustrating a focal displacement in curvature of the PCS and STS associated with hallucinations, that is strikingly similar to the sulcal kink previously observed in the STS of patients with hallucinations (21). With the local gyrification index (LGI) as a validated proxy for local sulcal shape, we exploited the sensitivity of structural covariance of functional RSNs. We hypothesized that deviations in frontal and temporal sulcal anatomy would reflect more general alterations in the topographical organization of large-scale networks; specifically, that patients with and without hallucinations would diverge in the inter-regional structural covariance within and between the salience and auditory networks. Consistent with the ubiquity and functional significance of asymmetries in the human brain (9, 27, 28), we replicated established sulcal lateralities and evaluated their significant association with hallucinations. Additionally, we complemented our analysis of cortical sulcation and gyrification with more usually studied metrics of cortical thickness and gray matter volume, which are plastic to aging, learning, and experience.

## Materials and methods

### Participants and study design

Two MRI datasets were re-purposed from independent studies of patients with recent-onset schizophrenia who underwent clinical assessment and 3T structural neuroimaging: (1) a predominantly White British sample assessed at multiple sites in the UK (29) and; (2) a Han Chinese sample assessed in Shanghai, China (30). See Supplemental Tables S1–S2 for the eligibility criteria and scanning sequence details. Patients were grouped into those with hallucinations (H+; n = 79 UK sample, n = 22 Shanghai sample) defined by a score > 2 on the PANSS P3 item for hallucinatory behaviour at the time of scanning, and those not experiencing hallucinations (H-; n = 43 UK sample, n = 37 Shanghai sample), as has been used previously (31). Additionally, 63 healthy controls (HC) were recruited into the Shanghai study. Studies were approved by the North West Manchester NHS Research Ethics Committee and Shanghai Mental Health Centre and the Institute of Psychology, respectively. Written, informed consent was obtained from all participants.

### MRI processing and measurement of sulcal patterns

Although primary sulci like the cingulate sulcus are relatively stable cortical landmarks across the population, secondary sulci, such as the PCS, exhibit substantial phenotypic complexity and inter-individual and inter-hemispheric variability (Figure 1), hindering techniques for their accurate identification and measurement. Paus and colleagues (32) proposed a qualitative 3-category classification of the PCS as prominent, present, or absent that has been widely deployed. However, ambiguities in PCS patterns pose boundary definition problems that obtained from all participants. introduce subjectivity into this classification scheme, and other nomenclatures and sulcal definitions have been proposed (33). Differences between measurement techniques, and even between experts using the same technique, are a cause of discrepancies in sulcal measurements (33). The current gold standard method for measuring the length of the PCS remains manual segmentation, a method that has proven sensitive to differences in hallucination occurrence (8). However, manual methods become increasingly intractable for large datasets and are unable to provide other biologically meaningful sulcal metrics, such as depth (23). Semi-automated methods show success in extracting detailed 3-dimensional sulcal curves, and even in identifying the primary sulci (34, 35), yet the open challenge lies in labeling variable sulci such as the PCS, which to date can only be completed with manual intervention. Accurate labeling of sulci is necessary to meaningfully determine the functional significance of sulcal architecture diversity in health and disease.

We used BrainVISA’s Morphologist 2015 pipeline (http://brainvisa.info/web/morphologist.html) to segment all cortical sulci. The superior temporal sulcus (STS) was automatically labeled with BrainVISA, while the paracingulate sulcus (PCS) was manually identified from whole-brain sulcal segmentations due to its high morphological variability. STS and PCS length and depth measurements in native participant space were calculated from the resultant labels. Length measurements were validated against manual measurements resulting in an intra-class correlation coefficient, ICC = 0.933 (95% CI: 0.922–0.941) for the left hemisphere and ICC = 0.943 (95% CI: 0.937–0.947) for the right. Details of the validation are given in Supplemental Figure S1 and Supplemental Tables S3–S4. To visualize the average morphology of the sulci of interest in each group (H+, H-, HC), we created 3D maps of the PCS and STS after linear spatial normalization to a common stereotactic space. To interrogate the locus of sulcal shifts, we applied the Human Connectome Project (HCP)-MMP1.0 multimodal surface-based anatomical atlas (36).

Cortical thickness (CT) and local gyrification index (LGI) were calculated using the FreeSurfer analysis package (v.6.0, http://surfer.nmr.mgh.harvard.edu/). Structural MRI data were analyzed with FSL-VBM (v.5.0.10, http://fsl.fmrib.ox.ac.uk/fsl), an optimized VBM protocol carried out with FSL tools.

### Replication and extension of sulcal and morphological asymmetries

The PCS has consistently been characterized to show a leftward asymmetry, wherein it is more often prominent or present in the left hemisphere than in the right (16, 26, 32). This leftward asymmetry is reduced in patients with schizophrenia (17–19). The superior temporal sulcus displays a robust rightward depth asymmetry at the base of Heschl’s gyrus, regardless of language dominance and certain atypical conditions (23). This asymmetry has not been investigated in schizophrenia, though reductions in volume, gyrification, and cortical thickness have all been observed in the superior temporal gyrus (5). Length and depth asymmetry indexes (AI) were computed for both PCS and STS: AI = 2*(R − L)/(R + L), for left (L) and right (R) measures, with a positive AI representing a longer or deeper sulci in the right hemisphere (Figure 4).

### Structural covariance networks for local gyrification index between and within auditory and salience networks

To succinctly estimate cortical topology over spatially extended RSNs, we computed the regional local gyrification index (LGI) as a proxy of sulcal morphology. The LGI is the ratio of the cortical area, including sulcal folds, to the cortical area visible on the pial surface. A higher LGI indicates a more involuted cortical surface, and is reduced by having fewer and shorter sulci. LGI is sensitive to cortical development (37), differentiates schizophrenia patients from healthy controls (38), and predicts transition to psychosis from clinical high-risk states (39). In our sample, the LGI across regions corresponding to the PCS and STS (Figure 5ab) was significantly correlated to both left and right PCS and STS length and depth (p< 0.05) (Supplemental Figure S3). We constructed inter-regional structural covariance matrices of LGI for 360 parcellated brain regions (180 per hemisphere) according to the HCP-MMP1.0 multimodal surface-based anatomical atlas (36) for each group: H+ n = 101; H- n = 80; HC n = 63, adjusted for age, sex, scanning site, and TIV. Matrices were re-organized into eight well-established and replicable resting-state networks (Supplemental Table S5) and LGI values between regions located within the same network were averaged, resulting in an 8×8 matrix for each group of network level LGI synchrony. (Figure 4, Supplemental Table S5). Following our hypotheses that deviations in sulcal anatomy would reflect alterations in the organization of specific RSNs, we evaluated the group differences in the inter-regional structural LGI covariance within and between the salience and auditory networks.

### Statistical analysis

Demographic and clinical differences between groups by sample (UK H+, UK H-, Shanghai H+, Shanghai H-, Shanghai HC) examined with one-way ANOVAs or chi-square tests for categorical data and t tests or Mann-Whitney U test for continuous variables, according to their distributions. To address hypotheses concerning PCS and STS length and depth differences, we performed per hemisphere linear regression analyses for PCS length by group, controlling for potential effects of age, sex, scanning center, and total intracranial volume (TIV). We performed this analysis in each dataset separately and in both combined. The homogeneity between UK and Shanghai samples and the ethnic invariance of sulcal morphology (26) enabled us to merge the datasets for subsequent analyses. Sulcal AIs were assessed within each group using one-sample t-tests and between groups (H+, H-, HC) using 1-way ANOVAs (Supplemental Table S6). To test the statistical significance between gyrification-based structural covariance networks, we performed nonparametric permutation testing with 5000 repetitions. For each iteration, the LGI parcellations of each participant were randomly assigned to one of three new groups with equivalent sample size to the original study groups (H+, H-, HC) and the between group difference in average correlation within and between the salience and auditory networks were calculated and plotted. This permutation approach preserves the LGI values and covariates for each participant, but shuffles group assignment across individuals. The observed differences in means were evaluated against the obtained permutation distributions, and a two-tailed p-value calculated based on its percentile position (< 5%). Resultant p-values were corrected by FDR (FDR< 0.05). To investigate the potential influence of laterality, we further decomposed the 8 networks into their hemisphere-specific component regions and repeated the analysis for the resultant 16×16 structural covariance matrices (Supplemental Table S7, Supplemental Figure S4). Analyses were conducted in RStudio (v.1.0.136) and Matlab (v.2017b). To reproduce prior cortical morphology analyses related to hallucinations (see (5) for a review), we conducted whole-brain analyses of cortical gyrification, thickness, and gray matter volume within each sample separately, corrected for multiple comparisons across each hemisphere with Monte Carlo simulation (10 000 iterations) and a cluster-forming threshold of p< 0.05 for surface-based analyses and nonparametric 2-sample t-tests using 5000 permutations and threshold-free cluster enhancement to identify areas in which grey matter volume differed between groups (p<0.05) within the medial prefrontal cortex.

## Results

### Multi-ethnic samples of schizophrenia patients with and without hallucinations, and healthy controls

Demographic and clinical differences between groups by sample (UK H+, UK H-, Shanghai H+, Shanghai H-, Shanghai HC) showed no significant group differences for gender, years of education, and olanzapine equivalent doses (Table 1). Post-hoc comparisons using Tukey HSD indicated no differences in mean age, but the 5-group 1-way ANOVA showed a significant main effect (F(4,236) = 2.44, p = 0.048). Individuals from the UK and Shanghai samples did not differ in their group-respective hallucination symptom scores (PANSS P3). However, the datasets differed in the ratio of patients with hallucinations to those without, with 65% of patients experiencing hallucinations in the UK sample, similar to that previously reported (22), but only 37% in the Shanghai sample. This could reflect recruitment differences between the two studies and/or cultural variation in the reporting or diagnosis of hallucinations (40). All participants in the UK sample and 93% of patients in the Shanghai sample were on stable antipsychotic medication at the time of scanning.

**Table 1.**
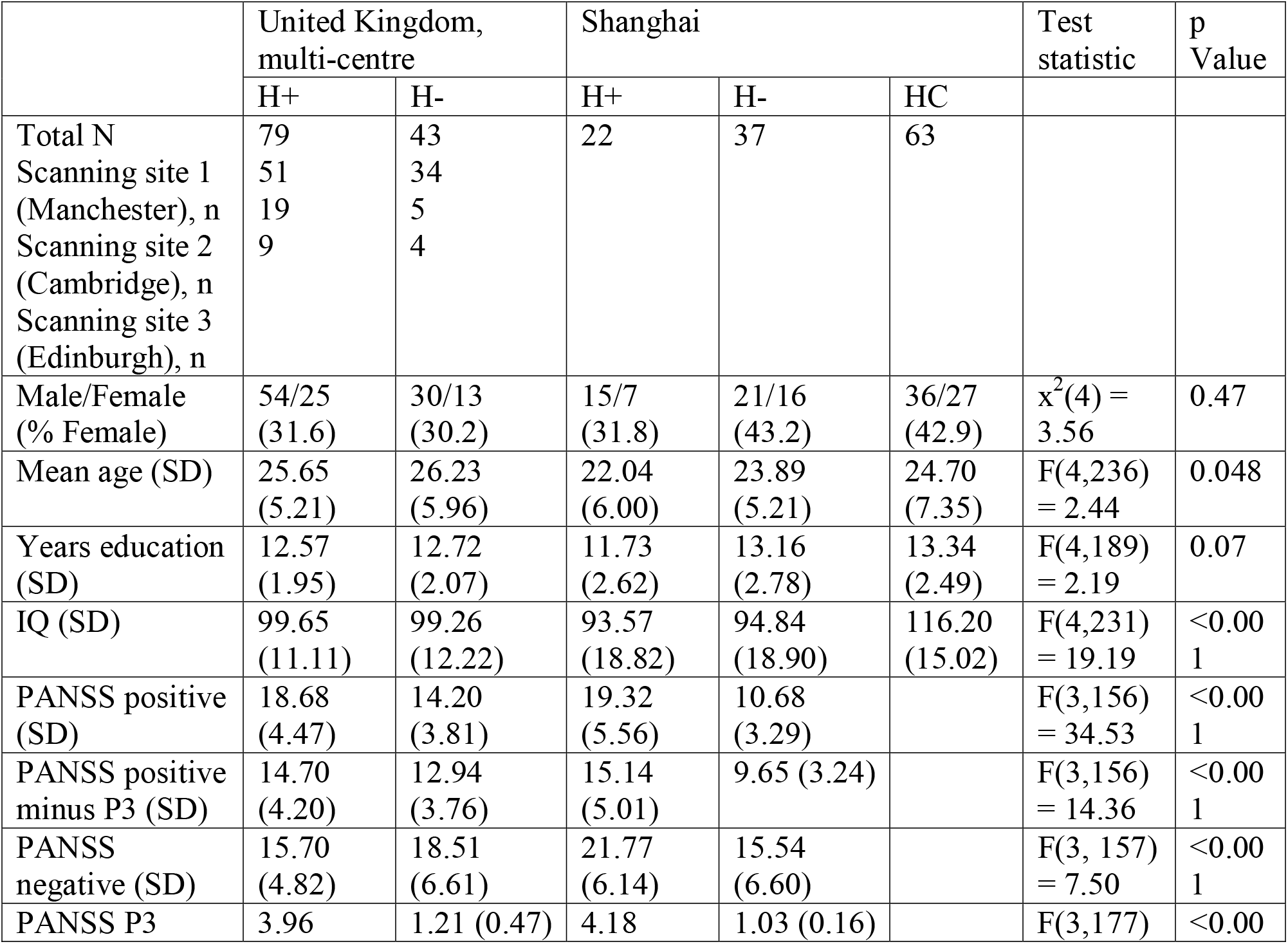
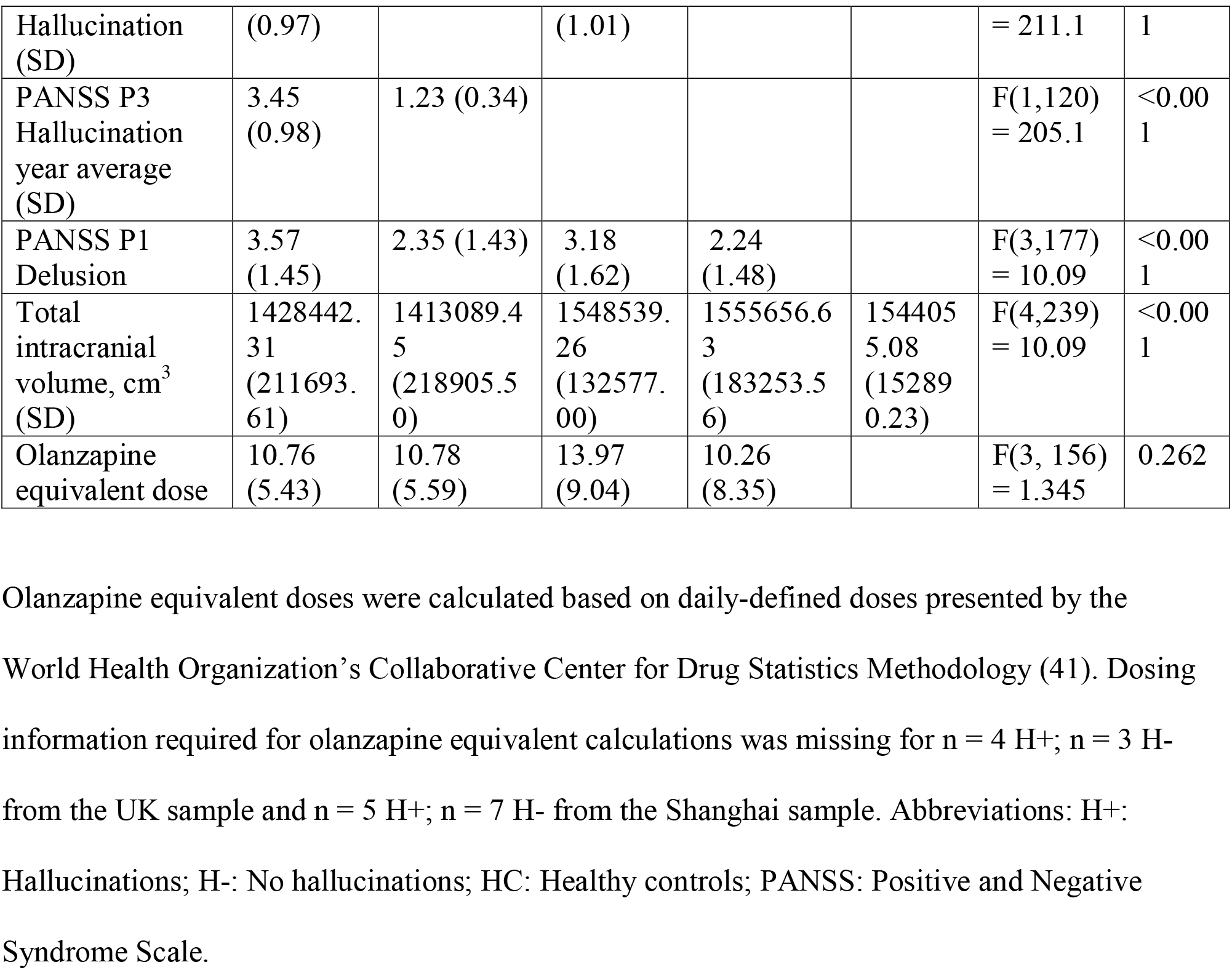
Characteristics of study participants.

### Left paracingulate sulcus length is reduced in schizophrenia patients with hallucinations in ethnically independent samples

We began by replicating, in independent samples involving a larger number of patients, our previous finding (8) that the length of the left hemisphere PCS calculated from structural MRI is reduced in schizophrenia patients with hallucinations compared to those without in both UK (t(114) = 2.293, p = 0.0237, β = 0.197) and Shanghai (t(109) = 2.332, p = 0.0215, β = 0.280), samples separately, and combined (t(227) = 3.264,p = 0.0013, β = 0.222). In the Shanghai sample, left PCS was also shorter in patients with hallucinations compared to healthy controls (t(109) = 2.716,p = 0.0077, β = 0.327) (Figure 2). These tests survived correction for additional covariates that were not included in the final model; i.e. years of education, IQ, global gyrification, total cortical surface area, olanzapine equivalent dose, PANSS P1 Delusion score, and PANSS positive symptoms minus P3 Hallucination score. There were no significant effects in the right hemispheric PCS (t(227) = −0.306, p = 0.760, β = −0.021), or in average PCS length (t(226) = 1.801,p = 0.0731, β = 0.121). The length of the right STS was reduced in patients with hallucinations compared to those without, but this reduction was not significant when controlling for age, sex, scanner site, and TIV t(227) = 1.240 p = 0.216, β = 0.090). Mean depth, however, was significantly reduced in the H+ group compared to HCs (t(227) = 2.381, p = 0.018, β = 0.209), and at trend for H- individuals (p = 0.098). The depth of the PCS was not significantly different between groups for either hemisphere. Bilateral PCS and STS length and depth measurements are reported in Supplemental Figure S2 and Supplemental Table S6.

**Figure 2.**
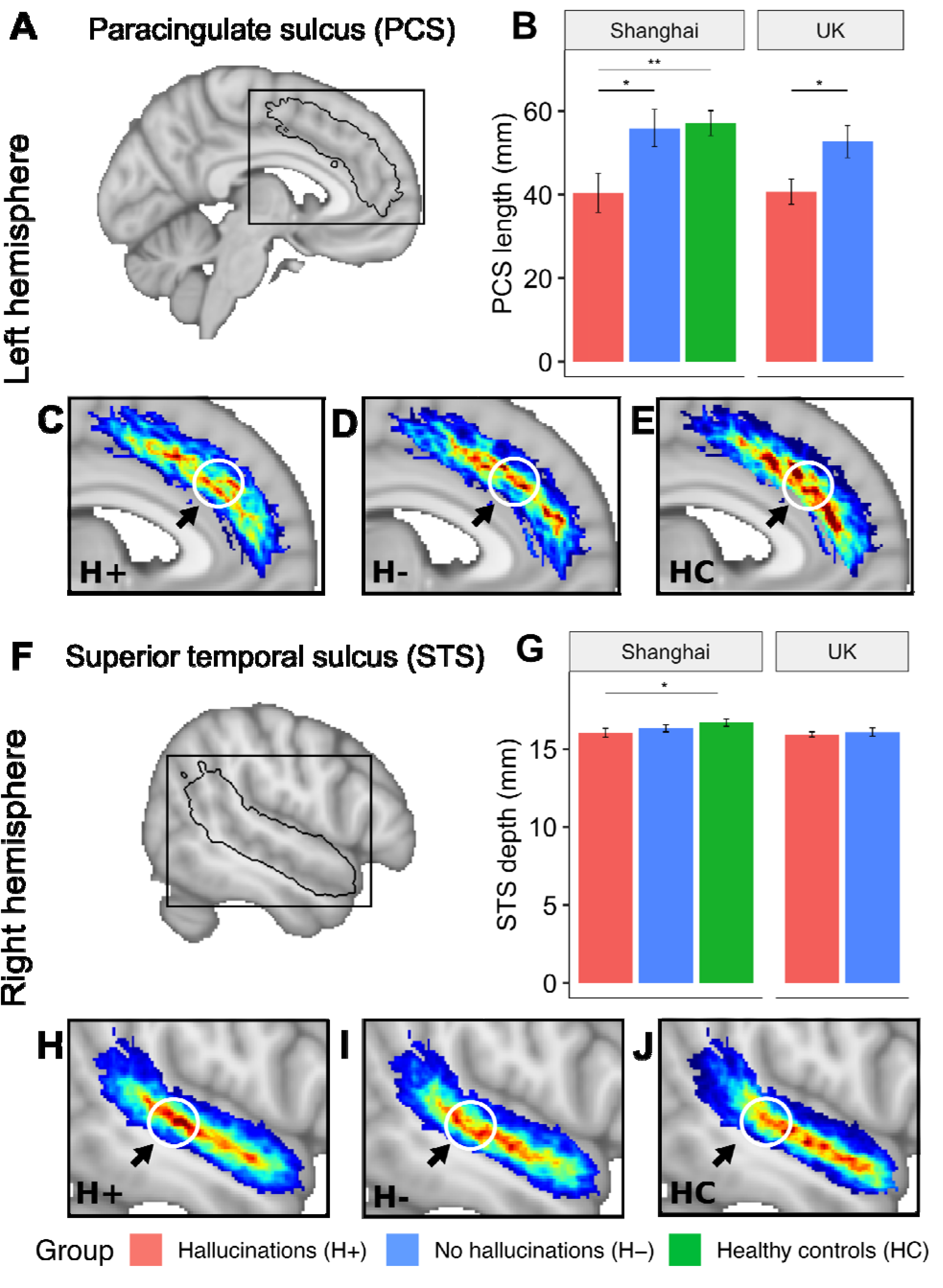
Local sulcal deviations associated with hallucinations. The length of the left PCS is significantly reduced in patients with hallucinations compared to those without and to HCs for both the Shanghai and UK multi-centre datasets (B). The mean depth of the right STS is significantly reduced in patients with hallucinations compared to HCs (G). Group-wise average sulcal maps after linear spatial normalization for (A) the left hemisphere paracingulate sulcus (PCS) (C–E) and (F) the right hemisphere superior temporal sulcus (STS) (H–J) display local curvature shifts between patients with (H+; n = 101) and without hallucinations (H-; n = 80) and healthy controls (HC; n = 63). This displacement makes the sulcus more direct and less arched for the H+ group and explains the reduced length of the left PCS. A parallel sulcal shift is present in the right STS. Arrows pointing to white circles indicate the area of maximum difference between the average sulcal maps of participants with and without hallucinations, which occurs in HCP area a32pr for the PCS and area STSdp for the STS. Average sulcal maps are linearly projected onto the MNI template. The colour bar represents the degree of overlap of sulci between participants in each group (H+, H-, HC), with red indicating higher overlap and thus the typical shape within group. Error bars denote the standard error of the mean. * p< 0.05; ** p< 0.01

### Reduced PCS length relates to a local displacement in curvature that emulates sulcal deviations in the STS

Although a shorter left PCS is a marker for hallucination status in patients with schizophrenia, the morphological features of the sulcus that lead to this observation have not previously been explored. We observed a focal displacement in the sulcal curvature such that the PCS of H+ participants was straighter, yet broken, in comparison to the more arched and continuous PCS characteristic of H- and HC participants (Figure 2). The PCS length difference is thus related to a difference in its curvature. In the right STS, a consistent pattern was observed similar to that detected by (21), with less arching in schizophrenia patients with hallucinations compared to those without. This observation is strikingly similar to the displacement of the sulcal junction between the right STS and its anterior branch in schizophrenia patients with hallucinations heard inside compared to outside the head (21). The left PCS shift mapped to area a32pr/d32 and right STS shift in STSdp (Figure 2). As both the anterior cingulate (42) and STS (43) show fine-grained gradients in functional organization, the detailed subdivisions of the HCP allowed us to interpret structural differences by their mapping to functional specializations. To increase the level of spatial detail within these focal and network-wide sulcal alterations, we visualized the correlations between left a32pr and right STSdp, the respective locations of the PCS and STS kinks, to all other parcellations (Figure 5).

**Figure 3.**
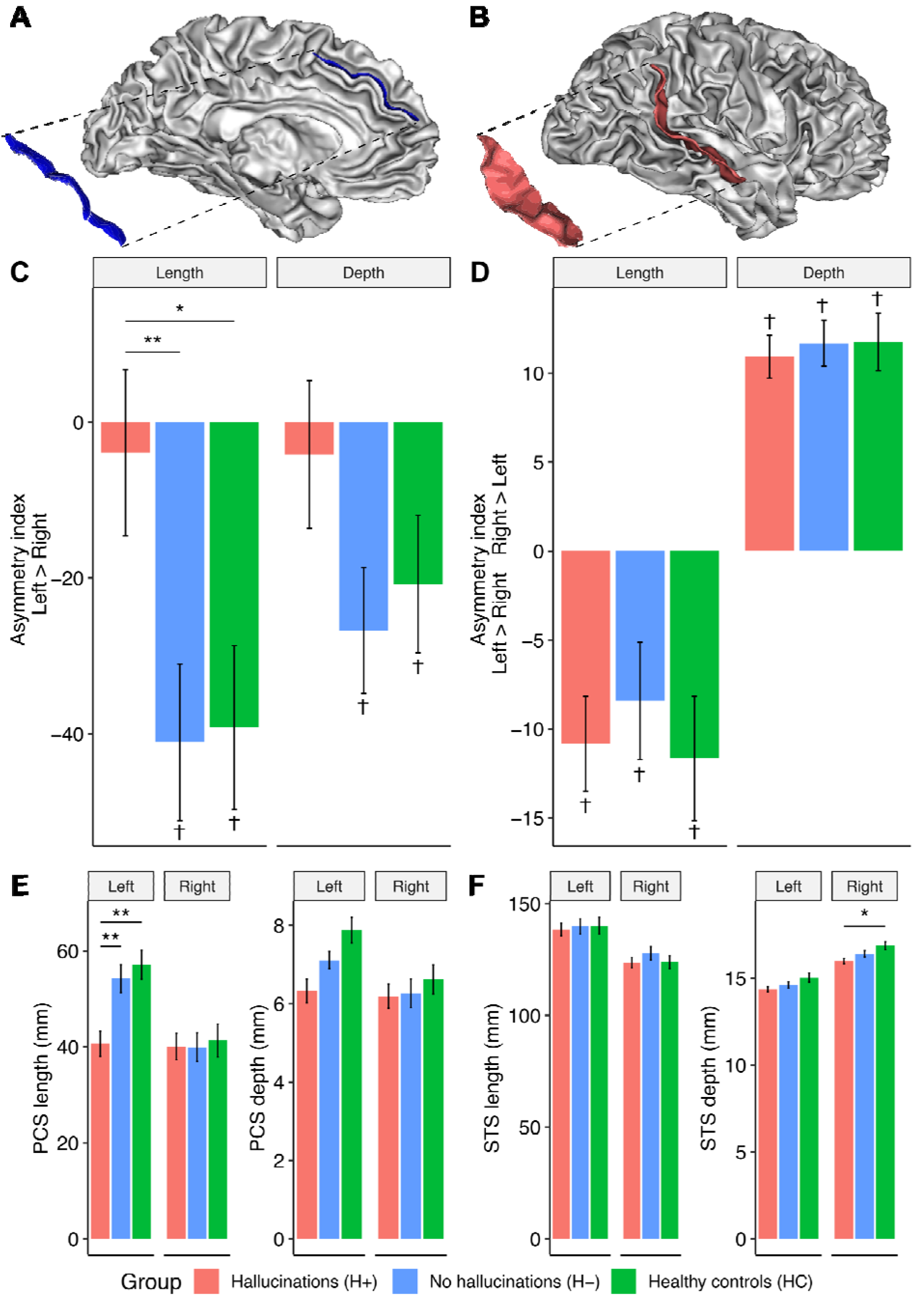
Asymmetry of length and depth measurements of the paracingulate and superior temporal sulci. 3D representations of the left hemisphere paracingulate sulcus (PCS) (A) and right hemisphere superior temporal sulcus (STS) (B). Asymmetry indices for the length and depth of the PCS (C) and STS (D). Positive values represent longer or deeper sulci in the right hemisphere compared to the left. Bilateral length and depth measurements for the PCS (E) and STS (F). Error bars denote the standard error of the mean. † Sulcal AI assessed within each group using 1-sample t-test. * Sulcal metrics assessed between groups using 1-way ANOVA for asymmetry indices and linear regression for hemisperic PCS and STS length and depth measurements, controlling for age, sex, scanner site, and total intracranial volume. * p< 0.05; ** p< 0.01.

### Replication and extension of sulcal and morphological asymmetries

One-sample t-tests revealed that the STS was significantly deeper in the right hemisphere than the left, but longer in the left hemisphere than the right, within all groups (p< 0.05). The PCS was significantly longer and deeper in the left hemisphere than the right for HCs and H- (p< 0.05), but the AI was not significant for H+. A 3-group 1-way ANOVA showed a significant main effect of group on PCS length AI (F(2,229) = 4.25,p = 0.0154). Post hoc comparisons using Tukey HSD revealed that the PCS length AI of patients with hallucinations was significantly reduced compared to those without (p = 0.0264), and trend level to HC (p = 0.0629), but was not different between HC and H- (p = 0.992). Previous reports of reduced PCS leftward asymmetry in schizophrenia, the majority in Western samples, thus appear to be likely driven by patients with hallucinations. AI for PCS depth and STS depth and length were not significant between groups.

### Structural covariance networks for local gyrification index show increased gyral synchrony between auditory and salience networks

Having established convergent sulcal deviations in the PCS and STS, we sought to investigate their covariance in the context of RSNs to gain insights into their developmental coordination, and how they might enable or reflect the emergence of functional networks that result in the experience of hallucinations. Structural covariance matrices organized by established large-scale functional resting-state networks (44) showed that mean (M) cross-correlation of the local gyrification index (LGI) for regions corresponding to the intersection of salience and auditory networks was significantly greater for H+ compared to H– individuals (M_H+_ = 0.501, M_H−_ = 0.355, M_HC_ = 0.375, q = 0.0084). The mean LGI cross-correlation was also significantly increased within each network (salience: M_H+_ = 0.493, M_H−_ = 0.371; M_HC_ = 0.375, q = 0.0147; auditory: M_H+_ = 0.631, M_H−_ = 0.532; M_HC_ = 0.523, q = 0.0292) after correcting for FDR < 0.05. Decomposing networks by hemisphere showed that these results were driven by increased mean LGI within the intrahemispheric right and inter- (lef-right) salience network, the intra-left auditory network, and between intra- and inter-salience and auditory networks (Supplemental Table S7, Supplemental Figure S4). When networks were decomposed by hemisphere, It is typical in structural covariance testing to discard low values of correlations between parcellated regions, attributing them to noise. Results were stable across a range of thresholds, but the greatest difference was observed with no threshold, suggesting that weak correlations, large in number, are important contributors to inter-regional cortical synchronization (Figure 4d). To increase the level of spatial detail within these focal and network-wide sulcal alterations, we visualized the correlations between the LGI in left a32pr and right STSdp, the respective locations of the PCS and STS kinks, to all other parcellations (Figure 5).

**Figure 4.**
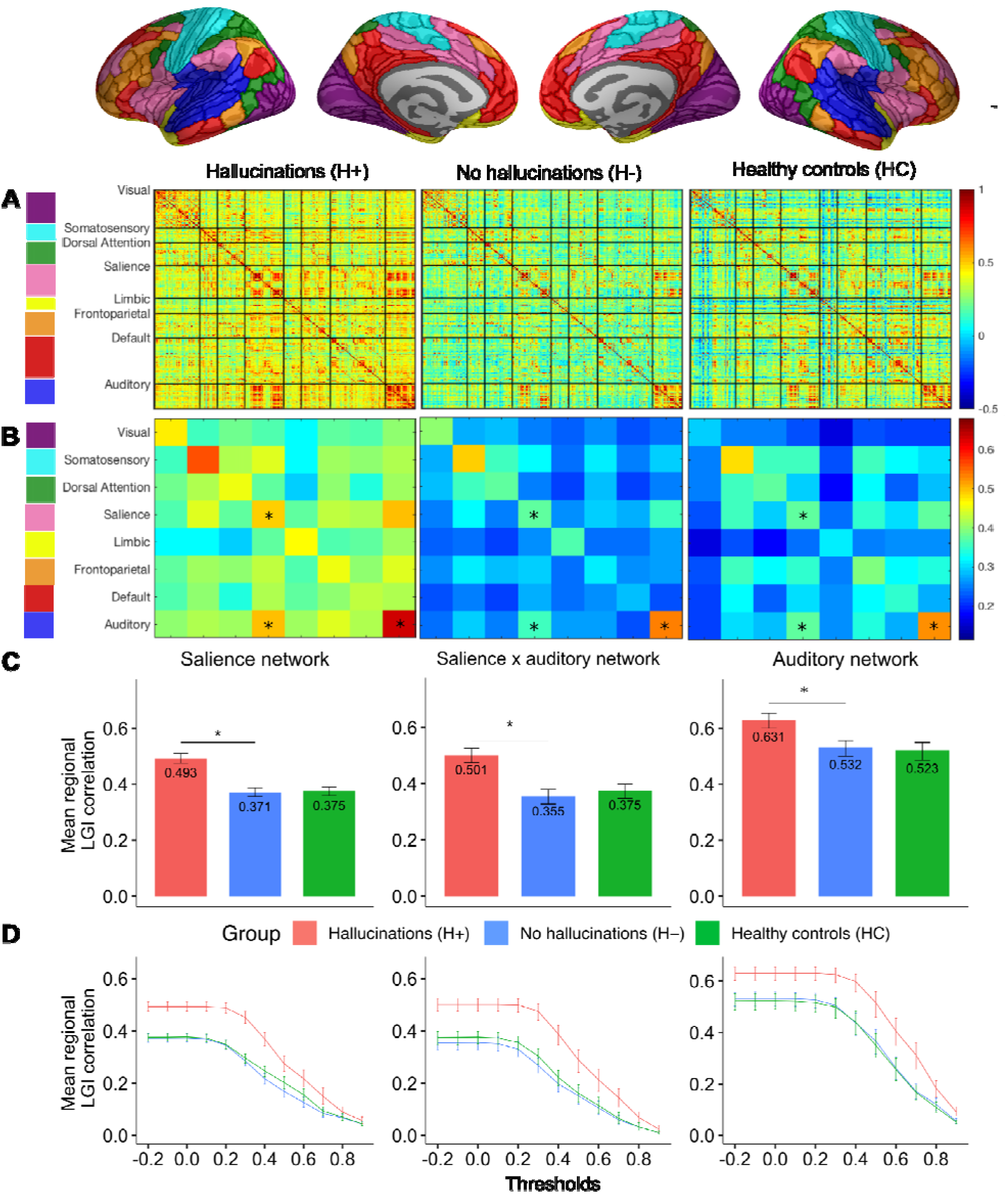
Stages in construction and analysis of gyrification-based correlation matrices. A. Local gyrification index (LGI) was computed for 360 parcellated brain regions (180 per hemisphere) according to the HCP-MMP1.0 atlas and was used to construct interregional Pearson’s correlation (360 × 360), adjusted for age, gender, scanning site, and intracranial volume for each of the 3 study groups (H+, H-, HC). Matrices were re-ordered according to 8 well-established and replicable resting-state networks. B. Each brain region was assigned a corresponding network and the LGI values between regions located within the same network were averaged, resulting in 8 × 8 group-wise matrices. C. Nonparametric permutation testing with 5000 resamplings was employed to test the significance of differences in patients with and without hallucinations in the mean LGI covariance within, and between, the salience and auditory networks. The observed differences in means were evaluated against the permutation null-distributions, and a two-tailed p-value calculated based on its percentile position (significance < 5%). D. Mean regional LGI partial correlation within brain regions corresponding to the salience, salience-auditory interaction, and auditory networks for correlation thresholds ranging from –0.2 – 1. Error bars represent the 95% bootstrap confidence intervals generated by resampling 5000 times with replacement across participants within each group. * FDR < 0.05.

**Figure 5.**
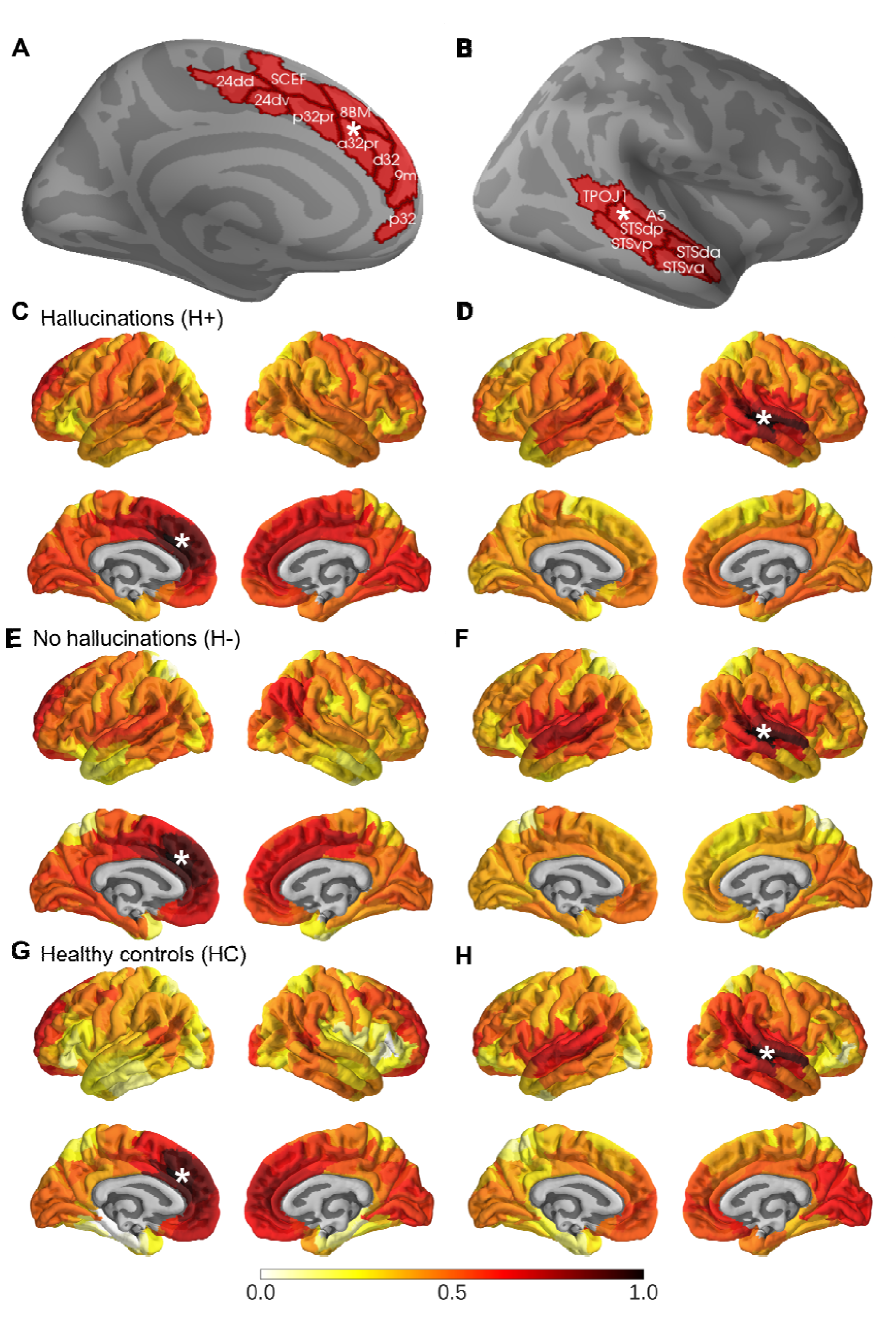
Whole-brain correlations for local gyrification index (LGI) in HCPMMP1.0-defined regions left a32pr and right STSdp. A. HCP regions that overlap with the paracingulate sulcus (PCS) (24dd, 24dv, SCEF, p32pr, a32pr, d32, p32, 8BM, 9m). B. HCP regions that overlap with the superior temporal sulcus (STS) (STSdp, STSda, STSvp, STSva, A5, TPOJ1). A white asterisk indicates the seed regions left a32pr and right STSdp, which were the loci of the sulcal displacements identified for the PCS and STS, respectively. C–H. Correlation between LGI in seed regions and all other 360 regions defined by the HCPMMP1.0 atlas for C– D. Hallucinations (H+; n = 101); E–F. No hallucinations (H-; n = 80); G–H. Healthy controls (HC; n = 63). The colour scale represents the correlation coefficient (Pearson’s *r*) of each regional LGI measure for the HCP parcellation to the LGI in left a32pr (C, E, H) and right STSdp (D, F, G).

Compared to patients without hallucinations, cortical statistical maps displayed decreased gyrification in H+ in the rostral middle frontal cortex and increased cortical thickness in the left inferior parietal and bilateral precuneus for the UK sample, and increased thickness in H+ for left lingual gyrus for both UK and Shanghai datasets (Supplemental Figure S5). There was greater gray matter volume in H+ across both datasets, as has been found previously (8), but the difference was only significant for the larger UK sample (Supplemental Table S8).

## Discussion

### Biological evidence for a perinatal structural network predisposing to hallucinations

Together, our findings suggest the existence of a discrete sulcal network that predisposes an individual to develop perceptual experiences untethered to reality. Bearing in mind that sulcal patterns are established during gestation and are fixed across the first decades of life, these results demonstrate an ethnically-invariant, postnatally-stable structural risk factor that is sensitive to subsequent life experiences and culturally-acquired expectations that color hallucination content. Hemispheric lateralization is a fundamental organizing axiom of the human brain that is apparent early in brain development and highly relevant for behaviour and cognition (27, 9, 28). We replicated documented sulcal asymmetries in the general population of a leftward length asymmetry for the PCS (17, 18, 19, 26) and rightward depth asymmetry for the STS (23) across two ethnically independent datasets. We found, however, an absence of the leftward PCS asymmetry in schizophrenia patients with hallucinations, due to a reduced length of the PCS compared to patients without hallucinations and to healthy controls. The rightward STS asymmetry was also lower, with a significantly diminished mean depth of the right STS in H+ patients compared to HCs. This supports empirical evidence that the reduction in structural and functional brain asymmetries in schizophrenia correlate with the severity of auditory hallucinations (28). Interestingly, certain genetic determinants of schizophrenia also modulate brain asymmetries in the auditory system, and it has recently been shown that there are lateralized genetic influences on frontal and temporal sulci (45), suggesting that sulcal descriptors offer insight into brain lateralization processes that relate to hallucinations in schizophrenia. The length and depth reductions observed here corresponded to distinct alterations in the sulcal geometry and increased LGI covariance within and between regions relating to salience and auditory networks, suggesting that perinatal alterations to the structural integrity of developing salience and auditory networks are related to experiencing hallucinations in early adulthood. Our results replicate and converge with previous research demonstrating differences in local folding patterns in cingulate and temporal regions (8, 20, 21, 38) and empirically integrates these spatially localized differences in support of hypotheses of aberrant fronto-temporal and salience network connectivity underpinning the experience of hallucinations in schizophrenia (12–14,46). We partially reproduce findings of abnormal surface-based morphology and gray matter volume related to hallucinations (5, 8). While these (non-sulcal) structural patterns were not wholly consistent between UK and Shanghai datasets, gray matter and cortical thickness are more subject to plastic changes as a function of aging, learning, experience, and medication, suggesting that sulcal topology presents a more robust and specific indicator of hallucinations. As sulcal patterns are established in utero and stable across the lifespan (6), these results pinpoint fetal brain features that predispose to for hallucinations in later illness, and may influence the way in which people experience their environment. The interaction between early neurodevelopment and experiential diversity helps explain, for instance, why trauma plays a major role in some hallucinations, but a minor or no role in others (47).

### Functional and behavioral implications of sulcal organization

We localized group differences in sulcal curvature to subdivisions of the STS and PCS. Each of these regions show heterogeneous functional specialization; the STS supports a range of social processes regionally specialized across a posterior-to-anterior axis, from language to theory of mind, to voice and face recognition (43), while the anterior cingulate encompasses heterogeneous cytoarchitecture, neurotransmitter receptors, and functional subdivisions that are associated with diverse cognitions of emotional reactivity, attention, fear, theory of mind, memory, and bodily sensations (42, 48). The sulcal kink identified in the STS mapped to STSdp, a region responsive to voice perception tasks and tasks engaging face and biological motion perception; and onto the border of midcingulate areas a32pr and d32 for the PCS. Area a32pr has functional connections primarily with other medial prefrontal regions, cingulate and insular regions, while d32 has additional functional connections to temporal and lateral parietal lobes (49). In rhesus monkeys, anterograde and retrograde tracing techniques have shown that despite regional heterogeneity in their projections, the medial prefrontal cortices are unified by bidirectional connections with the superior temporal cortex and neighboring auditory association cortices (50). One such connection, the arcuate fasciculus, shows reduced fractional anisotropy in the left hemisphere of schizophrenia patients with hallucinations compared to healthy controls (51), and left-lateralized reduced mean and radial diffusivity in children reporting psychotic experiences (52), suggesting a relationship between developmental changes to fronto-temporal association tracts and the increased LGI synchrony between salience and auditory regions observed here. We hypothesize that auditory hallucinations in schizophrenia patients, and the accompanying patterns of brain function, are due to differences in neuromaturational coupling between temporal and cingulate regions.

### Cortical folding mechanisms and functional-behavioral implications

What is the mechanism by which reduced morphological congruity between auditory and cingulate cortices develops and could it represent a structural presage of later-life hallucinations? Studies on the microstructural basis of sulcal macrostructure suggest that neuronal density changes, dendritic arborisation, synaptic pruning, and organization of axonal connections, individually or together, drive gyrification. A recent study using neurite orientation dispersion and density imaging, a novel multi-compartment diffusion MRI model of cortical microstructure, showed that neurite orientation dispersion is leftward asymmetric in frontal areas and rightward asymmetric in early auditory areas in two independent samples of healthy adults (53). The geometric effects of dendritic arborisation as a driver of cortical folding may therefore contribute to the sulcal asymmetries observed here. The evidence accumulating with regard to prenatal inflammatory exposure (54), maternal stress, obstetric complications (55), and expression of schizophrenia risk genes in fetal life as factors conferring vulnerability to schizophrenia would be of major relevance to how the intrauterine environment contributes to the variations in cortical folding that impact postnatal pathology.

Current theories of hallucinations in psychosis include those that propose perception is instantiated in a cortical hierarchy, mediated by feedforward and feedback processes that have layer-specific targets (56). Recently, layer-specific dynamics of neuronal activity by optogenetic stimulation have been shown to induce visual hallucinations in mice (56). That sulcal organization influences laminar gradations (57) links our findings to predictive processing accounts of perception and hallucinations (58). The increased cortical folding covariance between and within salience and auditory networks reported here could be a source, for instance, of the development of prior expectations with overly strong influence on perceptual inferences, or shifts in the weighting of external and internal information, such as inner speech. That cytoarchitecture relates to the distribution of neurotransmitter receptor binding sites and local concentrations of neurotransmitters and modulators is relevant in light of recent research linking dopamine release capacity and D2 receptor density to the severity of hallucinations in schizophrenia (59) and theories on the role of excitatory and inhibitory neurotransmitters in hallucinations (60).

### Ethnic invariance of hallucinations and opportunities for treatment

Hallucinations are a universal human experience, but are liable to local cultural variation. Whereas in Western countries voices are reported as commanding, violent and critical, and are attributed with diagnostic labels, in Eastern countries people are more likely to report rich relationships with their voices and ascribe positive meaning (61). Despite robust findings that ethnic milieu influences the content of hallucinations, no study has explored the impact of ethnic characteristics on the neuroanatomical basis for hallucinations. We show for the first time a common anatomical substrate associated with hallucinations in two ethnically distinct (British White vs. Han Chinese) groups of people with schizophrenia. As salience networks monitor the environment and directs attention, common anomalies to these networks in utero may interact with cultural expectations to influence how people attend to their environment (61), and experience hallucinations. Our findings hold clinical relevance. Although culture scaffolds perception, there is also evidence that the way in which people focus their attention on their environment can be changed through cultural priming, a technique to manipulate a person’s cultural value system (62). Although the mosaic of sulcal features of the brain are intransient and not appropriate targets for therapy, cultural priming of Eastern values proffers a novel opportunity for treatments to mitigate the distressing experience of hallucinations that is a common feature in Western cultures. At the very least, an individual’s cultural background should be considered in diagnosis and treatment.

### Limitations

The PANSS P3 measures current symptoms as opposed to lifetime history and does not distinguish between hallucination modality. The results could therefore be interpreted as a marker for treatment resistance, though the P3 score available for the UK dataset at 2, 4, 6, 9, and 12 months showed convergence with cross-sectional scores (Table 1). Our results may not be specific to auditory hallucinations, though these are the most common modality reported among schizophrenia patients. We additionally controlled for PANSS P1 Delusion score, and PANSS positive symptoms minus P3 Hallucination score for the sulcal length/depth analyses, which were not significant, indicating that our findings are specific to hallucinations, as opposed to overall psychosis severity. Though the two datasets were well matched for demographic and clinical characteristics, limited information was available for duration of untreated illness. The definition of the cortical folds in BrainVISA provides a stable and robust sulcal surface definition that is not affected by variations in cortical thickness or gray matter/white matter contrast (34), and is stable across the lifespan (6). Thus, we would not expect our results to be influenced by duration of illness or medication; in fact, medication did not differ between hallucinations status. However, longitudinal studies characterizing quantitative features of sulci linking structural development from fetal to adult life periods will be important in establishing whether the present risk factor has predictive value for conversion to psychosis or stratification for clinical interventions.

### Future questions, predictions and recommendations for research

Our findings of a sulcal network associated with hallucinations raise questions concerning their nosology and relationship to environmental factors. Do PCS-STS deviations selectively present in schizophrenia patients who develop hallucinations, or do they confer vulnerability to aberrant perceptions in other disorders, both with strong neurodevelopmental influences (ie. borderline personality disorder) and neurodegenerative causes (ie. Parkinson’s disease)? Are sulcal anomalies a modality-specific or general risk factor for hallucinations (63)? A recent study showing general sulcal deviations in schizophrenia patients with visual hallucinations suggests the latter (22). Does the prenatally formed sulcal pattern offer predictive value for identifying individuals who might develop hallucinations, for instance following bereavement or trauma, and are there any interventions that could mitigate the increased risk for hallucinations conferred by prenatally determined sulcal patterns?

## Conclusion

We progressed our previous work establishing the PCS as a marker for hallucinations and reality monitoring ability, embedding PCS anomalies into a broader neurodevelopmental framework of sulcal synchrony between the PCS-STS that confers vulnerability to develop hallucinations in people with a psychotic disorder, invariant to ethnic origin. We additionally recapitulated and enriched the biological detail of sulcal lateralities. These results suggest the existence of a network with in utero origins that acts as a universal structural marker for hallucinations in schizophrenia. Future work will investigate the ubiquity of these markers to other diagnoses and modalities of hallucinations on large datasets (64). Assessing brain development in the late fetal period and the interaction between sulcal architecture, functional connectivity and behavioral experiences will generate greater knowledge of the mechanisms supporting hallucinations, and holds synergistic prospects of novel treatment strategies and preventive interventions that target the salience and auditory networks. Our observations offer an interesting window to understanding normal perception, the boundaries between self and other, and the processes underlying how we construct our consciously experienced reality.

## Data Availability

The datasets reported in this manuscript are not publicly available because of lack of informed consent and ethical approval. Requests for sharing the anonymised datasets should be addressed to BD for the UK sample and RCKC for the Shanghai sample. Data analysis scripts and result files are available upon request, which should be addressed to the lead author.

## Acknowledgements

The authors were supported by grants and scholarships from the following sources: Gates Cambridge (CPER), National Key Research and Development Program (2016YFC0906402) (RCKC), National Science Fund China (81571317) (RCKC), the CAS Key Laboratory of Mental Health, Institute of Psychology (RCKC). Neuroimaging data was processed and archived on the University of Cambridge High Performance Hub for Clinical Informatics (HPHI), which was funded by part of the MRC Clinical Research Infrastructure Award (MR/M009041/1). The funders had no input in study design, data collection, data analysis, data interpretation, writing of the report, or the decision to submit for publication. The authors thank Aniket Patel, Rory Durham and Charlotte Jones for their contribution to validating the paracingulate sulcus manual tracing measurement protocol.

## Author contributions

CPER processed and analyzed all data, created the figures, and wrote the first draft of the manuscript. JS, GKM, JRG, and JSS conceived the project following research by JRG and JSS. JS and GKM directed the project and revised the manuscript. RCKC provided data for the Shanghai sample, and ZL, CY, and ZY contributed to its acquisition. JS, BD, and RU provided data for the UK sample and contributed to or supervised its acquisition. JS, GKM and JG supervised analysis. MA and AS contributed to processing data and validating manual and semi-automated methods. JY, DW, and PL offered technical support and contributed to methods development. AC provided technical guidance and offered theoretical input. All authors critically reviewed the manuscript and contributed to its writing and revision.

